# Learning and forecasting selection dynamics of SARS-CoV-2 variants from wastewater sequencing data using *Covvfit*

**DOI:** 10.1101/2025.03.25.25324639

**Authors:** David Dreifuss, Paweł Czyż, Niko Beerenwinkel

**Affiliations:** Department of Biosystems Science and Engineering, ETH Zurich, CH-4056 Basel, Switzerland; SIB Swiss Institute of Bioinformatics, CH-1015 Lausanne, Switzerland; ETH AI Center, ETH Zurich, CH-8092 Zurich, Switzerland

## Abstract

The COVID-19 pandemic has been driven by the emergence and spread of SARS-CoV-2 variants that confer a selective advantage over previously circulating strains. Estimating these selective advantages typically involves analyzing a large number of positive test samples through genomic sequencing. In this study, we present *Covvfit*, a statistical model and software package for estimating the fitness advantages of multiple competing variants using sequencing data derived from wastewater samples from different locations. We use our model to reconstruct the dynamics of variant competition across successive waves of the pandemic using over 5,000 samples from wastewater sequencing data collected between 2021 and 2025. We show through a comparison with clinical data that wastewater-based estimates of fitness advantages are efficient and accurate. Furthermore, we demonstrate that once variants surpass a low detection threshold, *Covvfit* can accurately predict their future dynamics over long prediction horizons.

## Introduction

Extensive sequencing efforts have led to the identification of thousands of SARS-CoV-2 variants from the genomic analysis of positive patient samples (*1*). While the large majority of these variants had little impact on the course of the pandemic, a few have been decisive in defining its trajectory. Some variants have shown increased transmissibility, while others have demonstrated the ability to evade immune response (*2, 3*). Variants with these traits possess a selective advantage, which can enable them to outcompete previously circulating strains, potentially triggering new waves of infections with severe public health consequences. Therefore, it is essential to estimate the selective advantage of arising genomic variants as accurately and as early as possible.

Large research efforts have demonstrated that it is possible to estimate the fitness advantage of variants using data obtained from clinical sequencing programs (*4*–*10*), and some of these methods have been integrated into epidemiological surveillance programs for risk assessment (*11*). Additionally, it has been shown that these models can accurately forecast the progression of the relative abundances of variants (*10*). However, these approaches depend on the availability of extensive clinical samples, which must be sequenced frequently and obtained through randomised sampling designs to avoid data collection bias. The collection of such data involves significant operational efforts and substantial costs, making it increasingly difficult to maintain. As a result, declining clinical sequencing efforts now provide insufficient data for rapid and accurate estimation of the fitness advantage of new SARS-CoV-2 variants.

During the COVID-19 pandemic, wastewater-based surveillance (or wastewater-based epidemiology, WBE) has become widespread and has proved to be a reliable data source to inform about the dynamics of SARS-CoV-2 and its variants. WBE offers several advantages over traditional clinical surveillance, including requiring orders of magnitude fewer samples and offering far less biased sampling schemes (*12*). Previous research has shown that logistic regression can be used to estimate the fitness of a single variant from wastewater next-generation sequencing (NGS) data (*13*–*15*) and from wastewater variant-specific PCR (*15, 16*). However, modern approaches for estimating fitness advantages from clinical sequencing data rely on multi-strain models that account for the competition between multiple co-circulating variants at once. A multi-strain statistical model designed for wastewater sequencing data is currently lacking, and there is no computational framework that allows for the systematic estimation and forecasting of variant dynamics from wastewater-based genomic surveillance.

Here, we propose *Covvfit*, a statistical model and software package designed to model the competition of SARS-CoV-2 variants using wastewater-derived sequencing data. Our method takes as input variant relative abundances computed by deconvolving wastewater sequencing using common wastewater NGS pipelines, and integrates data originating from different locations. *Covvfit* produces estimates of fitness advantages as well as forecasts of variant relative abundances together with uncertainty quantification in a time-efficient manner. We validate this approach using a genomic dataset of more than 5,000 wastewater samples collected across Switzerland between 2021 and 2025 (*13*). By comparing our estimates with those derived from clinical sequencing data, we demonstrate the strengths of wastewater-based epidemiology, including its scalability, cost-effectiveness, and ability to provide timely insights into variant dynamics and relative fitness. We additionally show that forecasts produced by *Covvfit* can be accurate up to prediction horizons of several months. Our findings underscores the potential of wastewater sequencing as a powerful tool for complementing traditional clinical surveillance in monitoring and forecasting the evolution of viral variants.

## Results

We developed *Covvfit*, a method to estimate the fitness advantages of SARS-CoV-2 variants using wastewater sequencing data. The method fits a multistrain selection model to time series data of variant relative abundances, which can be obtained by deconvolving sequenced wastewater samples using data de-mixing tools such as Freyja (*17*) or LolliPop (*18*). The package allows one to jointly model data collected at multiple locations (Figure 1a). By fitting the model, we estimate relative fitness values and obtain forecasts of future relative abundances of variants. To balance robustness, statistical efficiency, and scalability to large data sets, the method combines an evolutionary selection dynamics model with a quasi-likelihood approach (Methods).

**Fig. 1:**
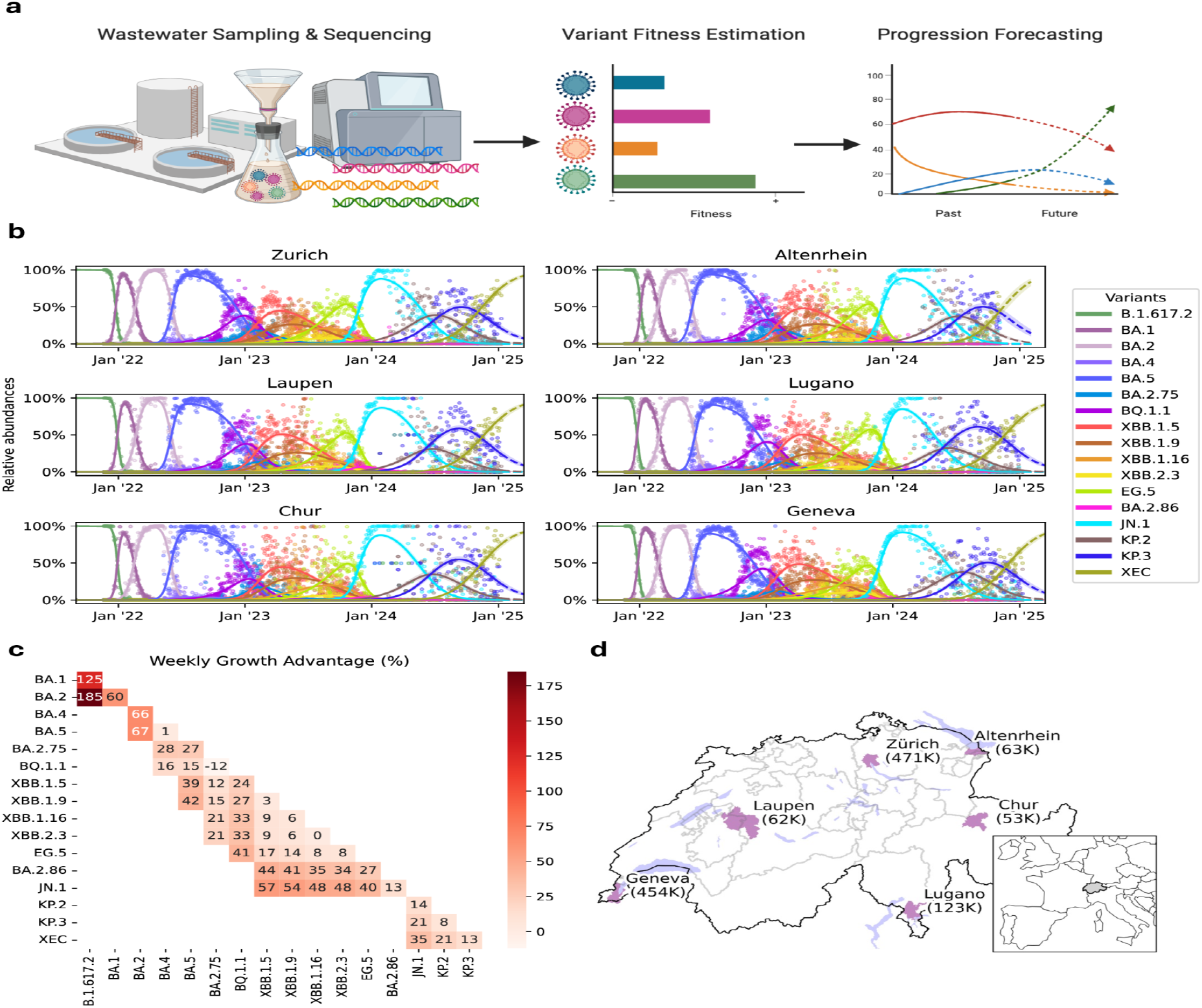
*Covvfit* estimates relative fitness of SARS-CoV-2 variants using wastewater sequencing data. **a** Overview of the procedure: wastewater sequencing data are deconvolved into variant relative abundance through time. *Covvfit* estimates the relative fitness advantages of different variants, which are used to produce forecasts of their relative abundances. **b** *Covvfit* model fitted to relative abundances estimated with *Lollipop* from wastewater NGS data from different locations in Switzerland. Dots are estimates of relative abundance for different variants on each day, lines are fitted model values and shaded bands are 95% confidence intervals for the fitted values. **c** Fitness advantages of variants in the rows relative to variants in the columns, for all pairs of variants having co-occurred. The fitness advantage is estimated by *Covvfit* using a generation time of 7 days. **d** Location of the treatment plants and sizes of the connected populations. Shaded regions represent catchment areas.

### *Covvfit* learns the fitness advantages of variants throughout the pandemic

We applied *Covvfit* to sequencing data from 5,266 Swiss wastewater samples collected from six locations between 2020 and 2025 (Figure 1d), tracking the successive introductions and spread of 18 main variants. *Covvfit* fitted the selection model to the 94,788 relative abundance data points in approximately 10 seconds on a standard laptop, reconstructing the variant dynamics over the course of the pandemic (Figure 1b). The method produced estimates and confidence intervals for the relative fitness of all co-circulating variants (Figure 1c, Supplementary Figure 2).

### *Covvfit* enables efficient early estimation of fitness advantages

To assess how early reliable estimates of variant fitness advantages can be obtained using *Covvfit*, we sequentially refitted the model using only data up to a given time and estimated the relative fitness advantage of emerging variants and their confidence intervals. We repeated this procedure over time to monitor how the estimates converged as more data was becoming available. For comparison, we performed this analysis for both wastewater sequencing data and all available clinical sequencing data from the regions in which the treatment plants are located.

### Early estimation of the fitness advantage of BA.2

We first focused on the introduction of the BA.2 variant, using samples collected from November 20, 2021, to February 28, 2022 (Figure 2 a). Fitness advantages estimated from wastewater sequencing data and from clinical sequencing data converged to similar values (Figure 2 c,e). Both estimates stabilized once BA.2 accounted for around 5% of cases in most surveyed WWTPs, and their rates of convergence were comparable. Notably, the wastewater-based estimates were based on orders of magnitude fewer samples (29 per week on average) as compared to clinical samples (approximately 1,250 per week on average; Figure 2 g). Importantly, sequencing clinical samples requires positive test results, such that orders of magnitude more clinical tests had to be performed. To further demonstrate the sample size requirements for estimating fitness advantages from clinical data, we subsampled the clinical samples to match sample sizes of the wastewater-derived data. The resulting fitness advantage estimates did not appear to converge (Supplementary Figure 1).

**Fig. 2:**
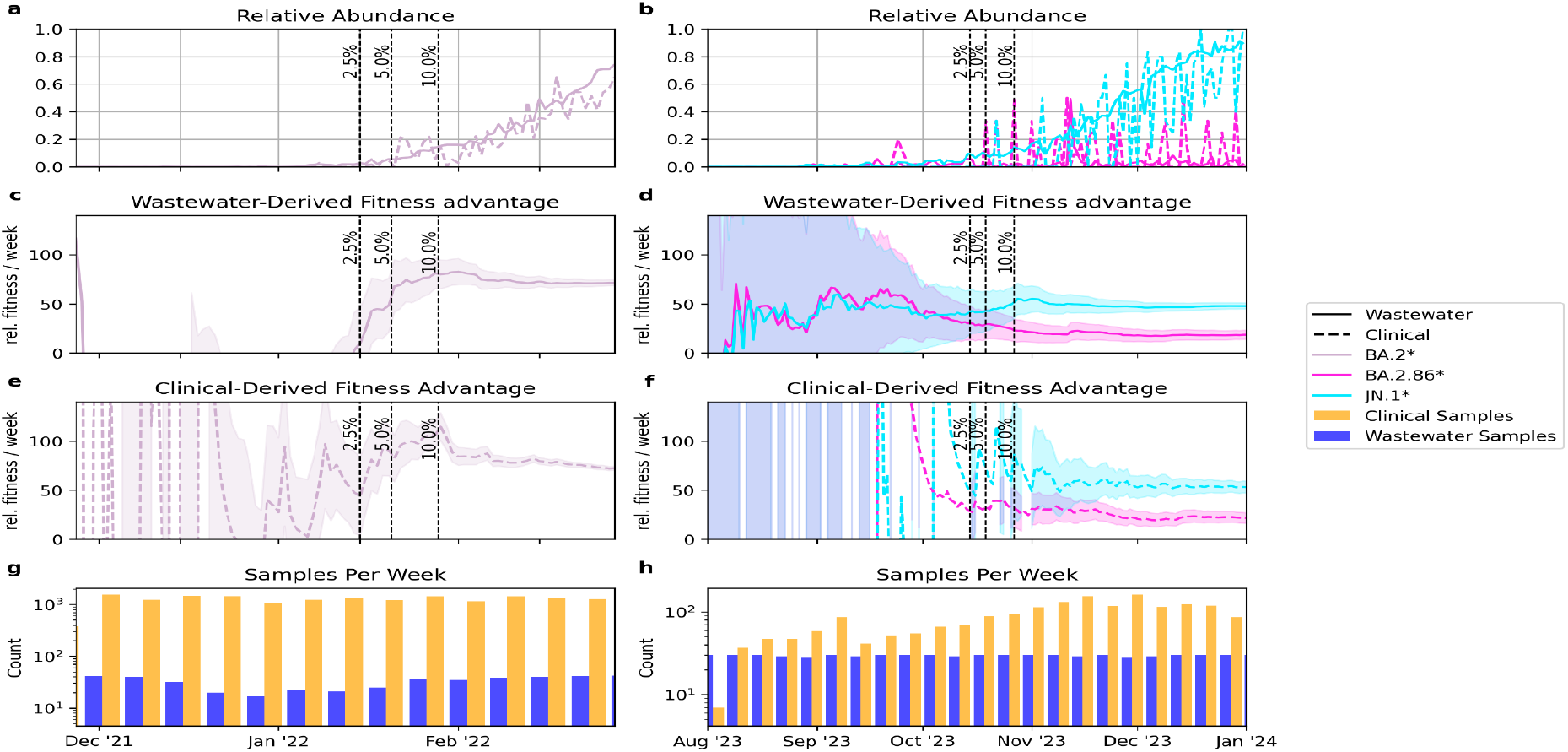
Early estimation of the fitness advantage of SARS-CoV-2 variants using *Covvfit*. **a** Introduction and spread of the BA.2 variant, based on relative abundance estimates from wastewater (solid line) and clinical (dashed line) sequencing data. For illustration, the time series is shown for the WWTP of Zurich and its catchment population. The vertical lines represent times at which BA.2 is estimated to cross the 2.5%, 5% and 10% relative abundance thresholds in most of the surveyed WWTPs. **b** Similar time series illustrating the introduction of the BA.2.86 and JN.1 variants. **c**,**d** Estimates of fitness advantage (lines) and 95% confidence intervals (shaded bands), computed in a joint model of 6 WWTPs using data available up to sequential timepoints. The fitness advantage for BA.2 is relative (in percent) to BA.1, and the fitness advantage for BA.2.86 and JN.1 is relative to EG.5. **e**,**f** Similar sequential estimation of selection, based on the clinical data available in the regions in which the WWTPs are located. **g**,**h** Number of samples per week used for the clinical data (blue) and for the wastewater data (orange). Importantly, clinical sequencing requires positive tests, such that the actual number of samples required is order of magnitudes higher.

### Early estimation of the fitness advantage of BA.2.86 and JN.1

We next investigated a scenario in which two variants were introduced at approximately the same time to assess how early we could determine which variant was fitter. Specifically, we examined the introduction of BA.2.86 and JN.1, using samples collected from October 21, 2022, to January 1, 2024 (Figure 2 b). As before, fitness advantages estimated from wastewater sequencing data and clinical sequencing data converged to similar values (Figure 2d, f). However, in this case, fewer clinical sequences were available (although still substantially more than wastewater samples, Figure 2 h), and estimates based on wastewater data converged faster than those based on clinical data. Wastewater data provided reliable estimates of fitness advantage early in the spread of BA.2.86 and JN.1. Importantly, wastewater-based estimates could infer early the fitness advantage of JN.1 over BA.2.86 (Supplementary Figure 5). Similarly as in the previous case, when we subsampled the clinical data to obtain a sample size comparable with the wastewater-derived data, the estimates of fitness became unstable (Supplementary Figure 1).

### *Covvfit* can accurately forecast future variant dynamics

To evaluate the predictive power of *Covvfit*, we tested its ability to forecast future variant dynamics. The experiment consisted of fitting the model to data available up to a given time and predicting the relative abundances of variants at future time points over different forecast horizons. To compute the prediction error, the model predictions were compared to a nonparametrically smoothed reference time series employing all the collected data.

### Forecasting during the transition from BA.1 to BA.2

We first studied the introduction of Omicron variant BA.1 and its replacement by BA.2 in early 2022 (Figure 3a). The prediction error shows that before enough data that includes BA.2 is available, forecasts are only accurate for horizons of several weeks (Figure 3c, 3e). The predictions for BA.1 at longer horizons exhibited a strong overestimation, while BA.2 was systematically underestimated (Figure 3g).

**Fig. 3:**
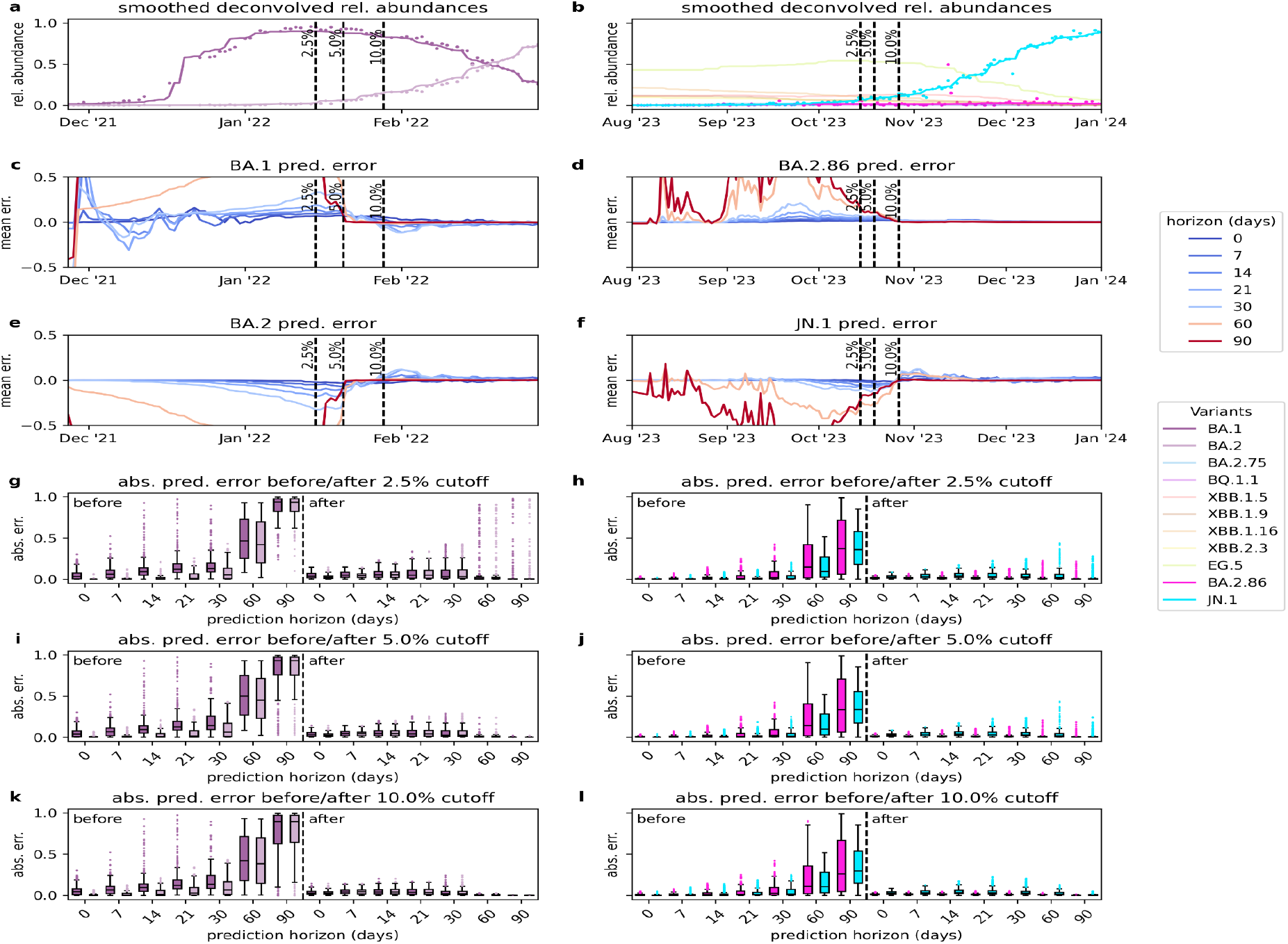
*Covvfit* forecasts prediction accuracy for relative abundances of variants in wastewater sequencing across two distinct time periods, namely the spread of BA.2 (left panels a, c, e, g, i, k) and the spread of BA.2.86 and JN.1 (right panels b, d, f, h, j, k). **a**,**b** Smoothed deconvolved relative abundances of different variants over time. For illustration, the time series is shown for the WWTP of Zurich. Dots represent raw data points, and solid lines indicate the running Gaussian kernel-weighted median, taken as baseline. Each variant is represented by a color. **c-f** Prediction error for the main variants for different forecast horizons (represented by different colors). The y-axis represents the mean prediction error (averaged over cities), where positive values indicate overestimation and negative values indicate underestimation. **g-l** Boxplots of absolute prediction errors, comparing errors for different variants and different horizon times (x-axis), before and after either BA.2 or JN.1 reaches 2.5%, 5%, or 10% relative abundance in most cities (marked with the vertical dashed lines). The central line within each box represents the median absolute error, while the box itself spans the interquartile range (IQR). The whiskers extend to the most extreme data points within 1.5 times the IQR, and outliers beyond this range are shown as individual dots. Each color corresponds to a different variant.

Once BA.2 crosses the 5% relative abundance threshold, the model learns that it has a substantial fitness advantage. Forecasts for horizons over 60 days become accurate, as the variant is correctly predicted to become dominant in the long run, while medium horizon forecasts retain some error as the dynamics during the displacement are still difficult to predict (Figure 3i). Once BA.2 crosses the 10% relative abundance threshold, the model corrects its predictions and forecasting dramatically improves, including for medium horizon forecasts. The error for both BA.1 and BA.2 drops significantly (Figure 3k).

### Simultaneous emergence of BA.2.86 and JN.1

A similar pattern is observed for the introduction of BA.2.86 and JN.1 in late 2023 (Figure 3b). Here, the model has to simultaneously learn the fitness of two new variants introduced at the same time, while in addition, there are more variants circulating in the background. In the early phase, before enough data was available, BA.2.86 is strongly overestimated for horizon times longer than several weeks, while JN.1 is underestimated (Figure 3d, 3f). This reflects that the model initially misattributed the dominant fitness advantage to BA.2.86 (Figure 2d, Supplementary Figure 5).

As more data becomes available and JN.1 reaches the 2.5% prevalence threshold, the model corrects its predictions and the error for both variants decreases dramatically, as the model correctly infers JN.1 as the fitter variant (Figure 2d, Supplementary Figure 5). Forecasts for horizons over 90 days are now accurate as the model predicts that JN.1 will become dominant while BA.2.86 will plummet (Figure 3h). Similar to the BA.2 case, once more data are available, the medium term dynamics of the variant relative abundances become predictable by the model and the forecast error is further reduced (Figure 3j,l).

## Discussion

On May 5, 2023, when the World Health Organization declared an end to the global public health emergency for COVID-19, it explicitly warned that the emergence of new variants continues to pose a threat to global health (*19*). Genomic surveillance remains crucial for tracking viral evolution and assessing public health risks. However, traditional clinical sequencing efforts have been significantly reduced worldwide, limiting our ability to assess and respond to emerging variants in a timely manner. Wastewater-based epidemiology can offer a scalable and cost-effective solution to this problem. In this study, we have introduced *Covvfit*, a statistical model and software package designed to estimate the fitness advantages and forecast the dynamics of SARS-CoV-2 variants using wastewater sequencing data.

Our findings show that *Covvfit* can accurately reconstruct the dynamics of variant competition throughout a pandemic. We showed that wastewater-based estimates of fitness can be reliably computed using orders of magnitude fewer samples than estimates based on clinical samples, and that they can accurately characterise the fitness of emerging variants early in their spread. As the fitness advantages of new variants indicate their potential to drive waves of infections and hospitalizations, early and accurate estimation is crucial for effective public health planning. Using wastewater samples, *Covvfit* offers a cost-effective and scalable alternative to clinical sequencing, making continuous public health planning informed by the fitness of emerging variants feasible even as clinical sequencing efforts decline.

We have shown that, given sufficient observation of all circulating variants, *Covvfit* can accurately forecast their future dynamics. However, when emerging variants still remain undetected or have not yet been introduced, predictions will necessarily be inaccurate. In the early stages of the spread of new variants, *Covvfit* rapidly learns the ranking of their fitnesses, allowing the public health researchers to predict which variants are likely to become dominant and which will go extinct. However, accurately forecasting their exact trajectories requires estimating not only their relative fitness ranks, but also quantifying their relative fitness values. As a result, sufficient data to obtain reliable quantitative medium-term forecasts becomes available later than for providing qualitative long-term predictions.

A major limitation of current wastewater-based variant surveillance approaches, which also applies here, is that deconvolution of wastewater sequencing data relies on variant definitions typically derived from clinical sequencing. Although some promising research has been devoted to learning variant profiles from wastewater data, they currently cannot fully replace clinically-derived variant definitions (*20, 21*). As a result, although wastewater surveillance is a powerful complementary tool, it cannot fully replace clinical sequencing for genomic epidemiology. Instead, these two approaches should be integrated into a synergistic surveillance framework: wastewater sequencing offers efficient and accurate population level monitoring and surveillance of variant dynamics, while clinical sequencing can enable precise characterization of newly emerging variants.

*Covvfit* shares several limitations common to models based on relative abundances of variants. The selection model attributes changes in observed abundances to the fitness values or statistical noise. The first consequence is that high rates of variant introduction can mimic selection effects in specific settings (*8*). This issue can be mitigated by analyzing multiple locations jointly. This will be more effective when considering distant locations, possibly through a global wastewater-based surveillance network. Second, the model is deterministic, so it may not be applicable to very small populations, where stochastic effects, such as genetic drift, play a large role (*22*). Third, *Covvfit* assumes fixed fitness values, which do not account for changes of the fitness landscape over time, such as, for example, shifts in the population immunity. However, previous analyses using clinical sequencing data have shown that predictive accuracy is not significantly affected by this assumption (*10*). Our results (Figure 3) agree with this finding, suggesting that the used model yields appropriate forecasting accuracy at a smaller computational cost than required by stochastic models. Finally, *Covvfit’s* predictions of future abundances are based on the assumption that no new variants will emerge in the considered prediction horizon, but unforeseeable introductions of new competitive variants will, in general, disrupt the dynamics learned on available data.

Beyond SARS-CoV-2, wastewater-based surveillance is increasingly being applied to other pathogens, many of which exhibit genomic diversity that is relevant to public health. For instance, influenza viruses are characterized by frequent genetic reassortment and antigenic drift, which can lead to the emergence of highly transmissible or immune-evasive strains (*23, 24*). However, for many other pathogens, we lack a detailed understanding of their genomic dynamics because they have not been studied at the same scale and level of detail as SARS-CoV-2 and influenza. *Covvfit* provides a general framework that could be tested to track variant competition in other pathogen populations, helping to identify competitive strains early and guide public health responses.

## Methods

### Selection model

The statistical model is composed of two components: the selection model, describing the dynamics of the competition between different variants and a statistical model for noise, linking the idealized abundances constructed by the model to the read count-based deconvoluted abundance values.

#### Single location

We consider *V* competing variants, indexed 1, …, *V*. At time *t*, the relative abundances of the variants are represented by a vector *p*(*t*) = (*p*_1_(*t*), *p*_2_(*t*), …, *p*_*V*_(*t*)). We assume survival-of-the-fittest selection dynamics (*22*), where the *v*-th variant has a fitness value *f*_*v*_, which is fixed in time, and the relative abundances change at a rate proportional to their fitness advantage over the average value:

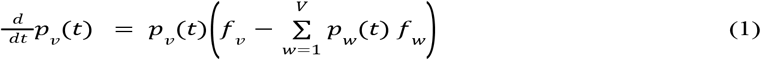

In this model, the fitness advantage of variant *v* over variant *w* is given by *s*_*vw*_= *f*_*v*_− *f*_*w*_. The model dynamics are determined only by the fitness advantages over a reference variant, *s*_*v*1_= *f*_*v*_− *f*_1_, rather than the fitness values *f*_*v*_, making the problem non-identifiable: adding the same constant to all fitness values does not change the dynamics of the model. Hence, without loss of generality, we set *f*_1_ = 0, to avoid this identifiability issue.

The fitness advantages in this parametrization represent relative rates per day, and can be converted to relative fitness by multiplying by the expected generation time *g*. Some studies prefer to report discrete time model fitness advantages, which can be obtained using the transformation *f* ↦ exp(*fg*) − 1 (*5*).

The set of ordinary differential equations (1) has the analytical solution

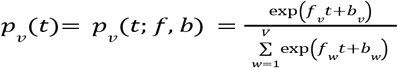

Where *b*_1_, *b*_2_, … *b*_*v*_ are constants given by the initial conditions. Note that adding a constant to all the values *b*_*v*_ does not change *p*_*v*_(*t*). Hence, similarly to constraining the fitness values, we constrain *b*_1_ = 0 for identifiability purposes.

#### Multiple locations

The model (1) above can be used to describe a collection of location-specific relative abundance vectors *p*_:*k*_ (*t*) = (*p*_1*k*_(*t*), …, *p*_*Vk*_(*t*)), one for each spatial location *k* ∈ {1,…, *K*} (e.g., the city or a district connected to one data collection system), such that *p*_*vk*_(*t*) is the relative abundance of variant *v* in location *k* at time *t*.

However, instead of fitting a collection of independent models, we describe a single model for multiple locations, in which a fixed fitness parameter *f*_*v*_ for each variant is shared between all locations. We expect that the introduction times of different variants at different locations may be different, which we accommodate by using location-specific parameters *b*_*vk*_:

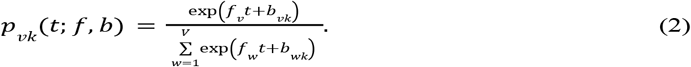

Similarly as above, we need to introduce identifiability constraints *f*_1_ = 0 for fitness values and *b*_1*k*_ = 0 for all locations *k* ∈ {1,…, *K*}. This results in a model with *N*_*param*_ = (*V* − 1) × *K* + (*V* − 1) free parameters.

### Statistical modeling of deconvoluted wastewater sequencing data

While the above mathematical model describes the behavior of the idealised abundance vectors *p*_:*k*_(*t*) at different locations *k* ∈ {1,…, *K*}, we cannot observe them directly. Instead, we use *Lollipop (18*) to deconvolute a wastewater sample to obtain an estimate of relative abundance vector collected at time point *t* and location *k*. Namely, let the obtained abundance vector be *y*_:*k*_(*t*) = (*y*_1*k*_(*t*), …, *y*_*Vk*_(*t*)), where *y*_*vk*_(*t*) represents the relative abundance of variant *v* ∈ {1, …, *V*} as obtained in the deconvolution procedure.

The observed value *y*_:*k*_(*t*) is connected to the idealized abundance *p*_:*k*_(*t*) by a complex underlying data-generating process, which consists of collecting a water sample with a small load of viral genome, amplifying it through next-generation sequencing method, mapping the obtained reads to reference genomes, and deconvolution to obtain the *y*_:*k*_(*t*) vector. Instead of framing this process as a generative model, we follow the quasi-likelihood approach popular in generalized linear modeling (*25, 26*).

We make the assumption that the deconvolved vector *y*_:*k*_(*t*) is a noisy, but unbiased, estimate of the idealised value *p*_:*k*_(*t*), that is *E*[*y*_:*k*_(*t*)] = _:*k*_(*t*). We also assume that the covariance *Cov*[*y*_:*k*_(*t*)] of the vector *y*_:*k*_(*t*) is given by a fixed matrix-valued covariance function *C* and a dispersion parameter *σ*^2^, such that

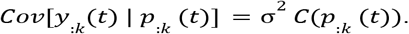

As *y*_*k*_(*t*) and *p*_*k*_(*t*) are probability vectors, we employ the quasi-multinomial covariance function, with the diagonal terms given by *C*(*p*_:*k*_)_*vv*_ = *p*_*vk*_(1 − *p*_*vk*_) and the off-diagonal terms given by *C*(*p*_:*k*_)_*vw*_=− *p*_*vk*_*p*_*wk*_. The dispersion parameter *σ*^2^ can then be estimated from the data to match the predicted covariance with the one observed in the data using a procedure we describe below.

The quasi-loglikelihood function corresponding to the quasi-multinomial model is given by

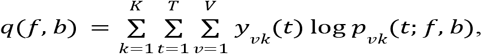

Where *p*_*vk*_(*t*; *f, b*) is given by (2). We numerically optimize the quasi-loglikelihood function to obtain the maximum quasi-likelihood estimate 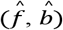. From this estimate, we further obtain estimates of the fitness advantages 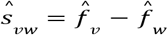 and predict the variant abundances 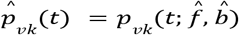 using formula (2).

Having fitted the model parameters, we estimate the dispersion parameter *σ*^2^ by matching the variance observed in the data to the one predicted by the model. Namely,

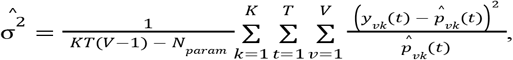

which estimates the dispersion parameter by using the generalized Pearson statistic calculated over the observed data points and adjusting the estimate by the number of free parameters *N*_*param*_ = (*V* − 1) × *K* + (*V* − 1) in the model(*25, 27*).

To robustify the computation of 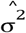 against outliers, we exclude from the summation elements where 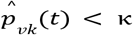 or 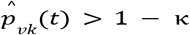, where κ = 0. 01 was chosen following perturbation experiments (Supplementary Figure 3).

The variance of the maximum-quasi-likelihood estimates is then approximated by 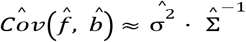. The matrix Σ is estimated via the observed Fisher information 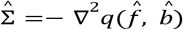. The confidence intervals for parameters are then computed using a normal approximation with covariance 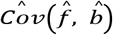. Confidence intervals for transformed parameters such as ŝ are computed using the delta method (*28*), by projecting the covariance using the Jacobian of the transformation. Confidence bands for the fitted or predicted values are computed on the logit scale (again, using the Delta method) before being back-transformed to the linear scale, to ensure a better coverage.

### Statistical model for clinical sequencing data

For the clinical sequencing data, we apply the same selection model as used for the wastewater data. However, instead of modeling the relative abundances derived from the deconvolution of wastewater sequencing data, we can directly count the clinical sequences assigned to different variants. At time *t*, in location *k*, the variant counts are denoted *z*_:*k*_(*t*) = (*z*_1*k*_(*t*), …, *z*_*Vk*_(*t*)). If we treat the sequences at a given location as independent realizations of a categorical random variable with probability vector *p*_:*k*_(*t*), the counts are distributed according to a multinomial distribution with probability vector *p*_:*k*_(*t*) and total sample size given by the number of sequences *n*_*k*_(*t*) for location *k* and time *t*. However, the assumption of different sequences from the same location being statistically independent is likely violated, due to non-random testing and sampling of tests. To take into account the added overdispersion, we again follow a quasi-likelihood approach. We assume that *E*[*z*_:*k*_(*t*)] = *n*_*k*_(*t*)*p*_:*k*_(*t*) and use a quasi-multinomial covariance function, such that the quasi-log-likelihood function is given by

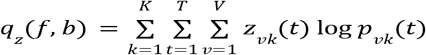

The rest of the inference follows similarly as for the wastewater data.

### Computational implementation

*Covvfit* is implemented in Python and leverages the automatic differentiation system of JAX for efficient optimization of the quasi-likelihood and accurate computsation of the observed Fisher information and Jacobians (*29*). This implementation ensures computational efficiency and precision, with the method scaling effortlessly to large surveillance projects. Additionally, *Covvfit* is designed for seamless interoperability with common wastewater genomic data processing pipelines using deconvolution tools such as *Lollipop (18*) and *Freyja (17*). The computational package is available in the Python Package Index (https://pypi.org/project/covvfit/), with its source code openly accessible at https://github.com/cbg-ethz/covvfit. The API documentation and tutorials are provided at https://cbg-ethz.github.io/covvfit/.

### Validation tests

#### Wastewater sequencing data

We used sequencing data from the Swiss wastewater monitoring program. The NGS raw data was processed with *V-pipe 3*.*0 (30*). In short, reads were filtered and trimmed with *PRINSEQ (31*) and aligned with *BWA-MEM (32*) to the reference sequence (GenBank accession number: NC_045512.2). We deconvolved mutation calls into relative abundances of selected variants using *Lollipop (18*), setting the kernel parameter to 0.1 to avoid smoothing.

We selected 5,590 samples from the WWTPs of Zurich (ZH), Altenrhein (SG), Laupen (BE), Lugano (TI), Chur (GR), and Geneva (GE) (Figure 1d) for the sampling period from 2021-05-01 to 2025-01-10. We discarded samples where the “undetermined” fraction of estimated variant relative abundances was higher than 0.01, which left 5,266 samples for analysis: 912 for Zurich (ZH), 879 for Altenrhein (SG), 864 for Laupen (BE), 881 for Lugano (TI), 912 for Chur (GR), and 818 for Geneva (GE).

We fitted the model jointly to the samples from all locations. To evaluate runtime, we recorded the total of CPU time plus system time during the actual fitting of the model.

#### Evaluating fitness estimates

We conducted a series of experiments. We fit the model using wastewater data using available data from time *t* to *t*′, and computed the relative fitness advantage of emerging variants, along with their confidence intervals. This process was repeated sequentially over time for varying *t*′ to assess how early reliable estimates could be obtained.

To assess the consistency between wastewater-derived estimates and estimates of selection based on clinical data, we conducted this analysis for both wastewater sequencing data and for clinical sequencing data from the regions in which the WWTPs are located. Clinical sequencing data was obtained from LAPIS (*33*), a public database of SARS-CoV-2 sequences. For each dataset, we fit the model to time series of variant relative abundances and computed the fitness advantages and confidence intervals over time.

The experiments were carried out over two periods, each capturing the emergence of key variants. The first period, from November 20, 2021, to February 28, 2022, covers the transition from BA.1 to BA.2. The second period, from October 21, 2022, to January 1, 2024, focuses on the emergence of BA.2.86 and JN.1. For both periods, the analysis was performed on six Swiss cities: Zurich, Altenrhein, Laupen, Lugano, Chur, and Geneva.

#### Evaluating predictions

To evaluate the accuracy of forecasts derived from the fitted model on wastewater data, we conducted a series of experiments. We first fitted the model using available data from time *t* to *t*′, then generated predictions for the relative abundances *y*^*pred*^(*t*′ + *h*) at different forecast horizons *h*. To mitigate the effects of sampling and deconvolution noise, we compared these predicted values to a smoothed time series *y*^*smooth*^(*t*′ + *h*) obtained using a Gaussian-weighted local median. This process was repeated for varying *t*′, and we recorded the prediction error.

The experiments were carried out on the same two time periods described above: November 20, 2021, to February 28, 2022, covering the emergence of BA.1 and BA.2, and October 21, 2022, to January 1, 2024, covering the emergence of BA.2.86, and JN.1. Analyses were performed by fitting the model jointly to the six Swiss cities. In both cases, the model predictions were evaluated at horizons up to 90 days, and the ground truth was smoothed using a Gaussian-weighted local median with a smoothing parameter of *σ*_*smooth*_ = 15. Accuracy was assessed by computing the mean prediction error over all cities for each investigated variant.

We further grouped errors based on whether they occurred before or after the latest rising variant crossed a 2.5%, 5% or 10% relative abundance threshold in most cities.

## Data Availability

The data and the code necessary to reproduce the results from this manuscript are available at https://github.com/cbg-ethz/covvfit/ and https://doi.org/10.5281/zenodo.15085753.

https://github.com/cbg-ethz/covvfit/

## Author contributions

Conceptualization: D.D, P.C, N.B. Methodology: D.D, P.C. Software: D.D, P.C. Validation: D.D, P.C. Formal analysis: D.D, P.C. Investigation: D.D, P.C. Resources: N.B. Data curation: D.D, P.C. Writing – original draft: D.D, P.C. Writing – review and editing: all authors. Visualization: D.D, P.C. Funding acquisition: D.D, N.B. Supervision: N.B.

## Data and materials availability

*Covvfit* code is available at https://github.com/cbg-ethz/covvfit/. The Supplementary Materials contain additional figures. The data and the code necessary to reproduce the results from this manuscript are available at https://doi.org/10.5281/zenodo.15085753.

## References

1. A. Rambaut, E. C. Holmes, Á. O’Toole, V. Hill, J. T. McCrone, C. Ruis, L. du Plessis, O. G. Pybus, A dynamic nomenclature proposal for SARS-CoV-2 lineages to assist genomic epidemiology. Nature Microbiology 5, 1403–1407 (2020).

2. P. V. Markov, M. Ghafari, M. Beer, K. Lythgoe, P. Simmonds, N. I. Stilianakis, A. Katzourakis, The evolution of SARS-CoV-2. Nature Reviews Microbiology 21, 361–379 (2023).

3. A. M. Carabelli, T. P. Peacock, L. G. Thorne, W. T. Harvey, J. Hughes, S. J. Peacock, W. S. Barclay, T. I. de Silva, G. J. Towers, D. L. Robertson, SARS-CoV-2 variant biology: immune escape, transmission and fitness. Nature Reviews Microbiology 21, 162–177 (2023).

4. H. S. Vöhringer, T. Sanderson, M. Sinnott, N. De Maio, T. Nguyen, R. Goater, F. Schwach, I. Harrison, J. Hellewell, C. V. Ariani, S. Gonçalves, D. K. Jackson, I. Johnston, A. W. Jung, C. Saint, J. Sillitoe, M. Suciu, N. Goldman, J. Panovska-Griffiths, E. Birney, E. Volz, S. Funk, D. Kwiatkowski, M. Chand, I. Martincorena, J. C. Barrett, M. Gerstung, Genomic reconstruction of the SARS-CoV-2 epidemic in England. Nature 600, 506–511 (2021).

5. C. Chen, S. A. Nadeau, I. Topolsky, M. Manceau, J. S. Huisman, K. P. Jablonski, L. Fuhrmann, D. Dreifuss, K. Jahn, C. Beckmann, M. Redondo, C. Noppen, L. Risch, M. Risch, N. Wohlwend, S. Kas, T. Bodmer, T. Roloff, M. Stange, A. Egli, I. Eckerle, L. Kaiser, R. Denes, M. Feldkamp, I. Nissen, N. Santacroce, E. Burcklen, C. Aquino, A. C. de Gouvea, M. D. Moccia, S. Grüter, T. Sykes, L. Opitz, G. White, L. Neff, D. Popovic, A. Patrignani, J. Tracy, R. Schlapbach, E. T. Dermitzakis, K. Harshman, I. Xenarios, H. Pegeot, L. Cerutti, D. Penet, A. Blin, M. Elies, C. L. Althaus, C. Beisel, N. Beerenwinkel, M. Ackermann, T. Stadler, Quantification of the spread of SARS-CoV-2 variant B.1.1.7 in Switzerland. Epidemics 37, 100480 (2021).

6. R. Viana, S. Moyo, D. G. Amoako, H. Tegally, C. Scheepers, C. L. Althaus, U. J. Anyaneji, P. A. Bester, M. F. Boni, M. Chand, W. T. Choga, R. Colquhoun, M. Davids, K. Deforche, D. Doolabh, L. du Plessis, S. Engelbrecht, J. Everatt, J. Giandhari, M. Giovanetti, D. Hardie, V. Hill, N.-Y. Hsiao, A. Iranzadeh, A. Ismail, C. Joseph, R. Joseph, L. Koopile, S. L. Kosakovsky Pond, M. U. G. Kraemer, L. Kuate-Lere, O. Laguda-Akingba, O. Lesetedi-Mafoko, R. J. Lessells, S. Lockman, A. G. Lucaci, A. Maharaj, B. Mahlangu, T. Maponga, K. Mahlakwane, Z. Makatini, G. Marais, D. Maruapula, K. Masupu, M. Matshaba, S. Mayaphi, N. Mbhele, M. B. Mbulawa, A. Mendes, K. Mlisana, A. Mnguni, T. Mohale, M. Moir, K. Moruisi, M. Mosepele, G. Motsatsi, M. S. Motswaledi, T. Mphoyakgosi, N. Msomi, P. N. Mwangi, Y. Naidoo, N. Ntuli, M. Nyaga, L. Olubayo, S. Pillay, B. Radibe, Y. Ramphal, U. Ramphal, J. E. San, L. Scott, R. Shapiro, L. Singh, P. Smith-Lawrence, W. Stevens, A. Strydom, K. Subramoney, N. Tebeila, D. Tshiabuila, J. Tsui, S. van Wyk, S. Weaver, C. K. Wibmer, E. Wilkinson, N. Wolter, A. E. Zarebski, B. Zuze, D. Goedhals, W. Preiser, F. Treurnicht, M. Venter, C. Williamson, O. G. Pybus, J. Bhiman, A. Glass, D. P. Martin, A. Rambaut, S. Gaseitsiwe, A. von Gottberg, T. de Oliveira, Rapid epidemic expansion of the SARS-CoV-2 Omicron variant in southern Africa. Nature 603, 679–686 (2022).

7. C. H. van Dorp, E. E. Goldberg, N. Hengartner, R. Ke, E. O. Romero-Severson, Estimating the strength of selection for new SARS-CoV-2 variants. Nature Communications 12, 1–13 (2021).

8. E. Volz, Fitness, growth and transmissibility of SARS-CoV-2 genetic variants. Nature Reviews Genetics 24, 724–734 (2023).

9. E. Volz, S. Mishra, M. Chand, J. C. Barrett, R. Johnson, L. Geidelberg, W. R. Hinsley, D. J. Laydon, G. Dabrera, Á. O’Toole, R. Amato, M. Ragonnet-Cronin, I. Harrison, B. Jackson, C. V. Ariani, O. Boyd, N. J. Loman, J. T. McCrone, S. Gonçalves, D. Jorgensen, R. Myers, V. Hill, D. K. Jackson, K. Gaythorpe, N. Groves, J. Sillitoe, D. P. Kwiatkowski, S. Flaxman, O. Ratmann, S. Bhatt, S. Hopkins, A. Gandy, A. Rambaut, N. M. Ferguson, Assessing transmissibility of SARS-CoV-2 lineage B.1.1.7 in England. Nature 593, 266–269 (2021).

10. E. Abousamra, M. Figgins, T. Bedford, Fitness models provide accurate short-term forecasts of SARS-CoV-2 variant frequency. PLOS Computational Biology 20, e1012443 (2024).

11. Technical briefing, SARS-CoV-2 variants of concern and variants under investigation in England. https://assets.publishing.service.gov.uk/media/61c5a722e90e071962ef0eae/technical-briefing-33.pdf.

12. Website. https://journals.asm.org/doi/10.1128/cmr.00103-22.

13. K. Jahn, D. Dreifuss, I. Topolsky, A. Kull, P. Ganesanandamoorthy, X. Fernandez-Cassi, C. Bänziger, A. J. Devaux, E. Stachler, L. Caduff, F. Cariti, A. T. Corzón, L. Fuhrmann, C. Chen, K. P. Jablonski, S. Nadeau, M. Feldkamp, C. Beisel, C. Aquino, T. Stadler, C. Ort, T. Kohn, T. R. Julian, N. Beerenwinkel, Early detection and surveillance of SARS-CoV-2 genomic variants in wastewater using COJAC. Nat Microbiol 7, 1151–1160 (2022).

14. F. S. Brunner, A. Payne, E. Cairns, G. Airey, R. Gregory, N. D. Pickwell, M. Wilson, M. Carlile, N. Holmes, V. Hill, H. Child, J. Tomlinson, S. Ahmed, H. Denise, W. Rowe, J. Frazer, R. van Aerle, N. Evens, J. Porter, COVID-19 Genomics UK (COG-UK) Consortium, K. Templeton, A. R. Jeffries, M. Loose, S. Paterson, Utility of wastewater genomic surveillance compared to clinical surveillance to track the spread of the SARS-CoV-2 Omicron variant across England. Water Res. 247, 120804 (2023).

15. D. Dreifuss, J. S. Huisman, J. C. Rusch, L. Caduff, P. Ganesanandamoorthy, A. J. Devaux, C. Gan, T. Stadler, T. Kohn, C. Ort, N. Beerenwinkel, T. R. Julian, Estimated transmission dynamics of SARS-CoV-2 variants from wastewater are robust to differential shedding, bioRxiv (2023)p. 2023.10.25.23297539.

16. L. Caduff, D. Dreifuss, T. Schindler, A. J. Devaux, P. Ganesanandamoorthy, A. Kull, E. Stachler, X. Fernandez-Cassi, N. Beerenwinkel, T. Kohn, C. Ort, T. R. Julian, Inferring transmission fitness advantage of SARS-CoV-2 variants of concern from wastewater samples using digital PCR, Switzerland, December 2020 through March 2021. Euro Surveill. 27 (2022).

17. S. Karthikeyan, J. I. Levy, P. De Hoff, G. Humphrey, A. Birmingham, K. Jepsen, S. Farmer, H. M. Tubb, T. Valles, C. E. Tribelhorn, R. Tsai, S. Aigner, S. Sathe, N. Moshiri, B. Henson, A. M. Mark, A. Hakim, N. A. Baer, T. Barber, P. Belda-Ferre, M. Chacón, W. Cheung, E. S. Cresini, E. R. Eisner, A. L. Lastrella, E. S. Lawrence, C. A. Marotz, T. T. Ngo, T. Ostrander, A. Plascencia, R. A. Salido, P. Seaver, E. W. Smoot, D. McDonald, R. M. Neuhard, A. L. Scioscia, A. M. Satterlund, E. H. Simmons, D. B. Abelman, D. Brenner, J. C. Bruner, A. Buckley, M. Ellison, J. Gattas, S. L. Gonias, M. Hale, F. Hawkins, L. Ikeda, H. Jhaveri, T. Johnson, V. Kellen, B. Kremer, G. Matthews, R. W. McLawhon, P. Ouillet, D. Park, A. Pradenas, S. Reed, L. Riggs, A. Sanders, B. Sollenberger, A. Song, B. White, T. Winbush, C. M. Aceves, C. Anderson, K. Gangavarapu, E. Hufbauer, E. Kurzban, J. Lee, N. L. Matteson, E. Parker, S. A. Perkins, K. S. Ramesh, R. Robles-Sikisaka, M. A. Schwab, E. Spencer, S. Wohl, L. Nicholson, I. H. McHardy, D. P. Dimmock, C. A. Hobbs, O. Bakhtar, A. Harding, A. Mendoza, A. Bolze, D. Becker, E. T. Cirulli, M. Isaksson, K. M. Schiabor Barrett, N. L. Washington, J. D. Malone, A. M. Schafer, N. Gurfield, S. Stous, R. Fielding-Miller, R. S. Garfein, T. Gaines, C. Anderson, N. K. Martin, R. Schooley, B. Austin, D. R. MacCannell, S. F. Kingsmore, W. Lee, S. Shah, E. McDonald, A. T. Yu, M. Zeller, K. M. Fisch, C. Longhurst, P. Maysent, D. Pride, P. K. Khosla, L. C. Laurent, G. W. Yeo, K. G. Andersen, R. Knight, Wastewater sequencing reveals early cryptic SARS-CoV-2 variant transmission. Nature 609, 101–108 (2022).

18. D. Dreifuss, I. Topolsky, P. Icer Baykal, N. Beerenwinkel, Tracking SARS-CoV-2 genomic variants in wastewater sequencing data withLolliPop, bioRxiv (2022). 10.1101/2022.11.02.22281825.

19. WHO Director-General’s opening remarks at the media briefing – 5 May 2023. https://www.who.int/director-general/speeches/detail/who-director-general-s-opening-remarks-at-the-media-briefing 5-may-2023.

20. I. Ellmen, A. K. Overton, J. J. Knapp, D. Nash, H. Ho, Y. Hungwe, S. Prasla, J. I. Nissimov, T. C. Charles, Reconstructing SARS-CoV-2 lineages from mixed wastewater sequencing data. Sci. Rep. 14, 20273 (2024).

21. L. Fuhrmann, B. Langer, I. Topolsky, N. Beerenwinkel, VILOCA: sequencing quality-aware viral haplotype reconstruction and mutation calling for short-read and long-read data. NAR Genom. Bioinform. 6, lqae152 (2024).

22. M. A. Nowak, Evolutionary Dynamics: Exploring the Equations of Life (Belknap Press, London, England, 2006).

23. R. Chen, E. C. Holmes, The evolutionary dynamics of human influenza B virus. J Mol Evol 66, 655–663 (2008).

24. J. Steel, A. C. Lowen, Influenza A virus reassortment. Curr Top Microbiol Immunol 385, 377–401 (2014).

25. P. McCullagh, J. A. Nelder, Generalized Linear Models (Routledge, 2nd Edition., 2019).

26. R. W. M. Wedderburn, Quasi-likelihood functions, generalized linear models, and the Gauss—Newton method. Biometrika 61, 439–447 (1974).

27. A. R. Linero, Bayesian Nonparametric Quasi Likelihood, arXiv [stat.ME] (2024). http://arxiv.org/abs/2405.20601.

28. L. Held, D.S. Bové, Applied Statistical Inference: Likelihood and Bayes (Springer Science & Business Media, 2013).

29. Jax: Composable Transformations of Python+NumPy Programs: Differentiate, Vectorize, JIT to GPU/TPU, and More (Github; https://github.com/jax-ml/jax).

30. L. Fuhrmann, K. P. Jablonski, I. Topolsky, A. A. Batavia, N. Borgsmüller, P. I. Baykal, M. Carrara, C. Chen, A. Dondi, M. Dragan, D. Dreifuss, A. John, B. Langer, M. Okoniewski, L. du Plessis, U. Schmitt, F. Singer, T. Stadler, N. Beerenwinkel, V-pipe 3.0: a sustainable pipeline for within-sample viral genetic diversity estimation. Gigascience 13 (2024).

31. R. Schmieder, R. Edwards, Quality control and preprocessing of metagenomic datasets. Bioinformatics 27, 863–864 (2011).

32. H. Li, R. Durbin, Fast and accurate short read alignment with Burrows-Wheeler transform. Bioinformatics 25, 1754–1760 (2009).

33. C. Chen, A. Taepper, F. Engelniederhammer, J. Kellerer, C. Roemer, T. Stadler, LAPIS is a fast web API for massive open virus sequencing data. BMC Bioinformatics 24, 1–13 (2023).

