## Supplementary figures for "Learning and forecasting selection dynamics of SARS-CoV-2 variants from wastewater sequencing data using *Covvfit*"

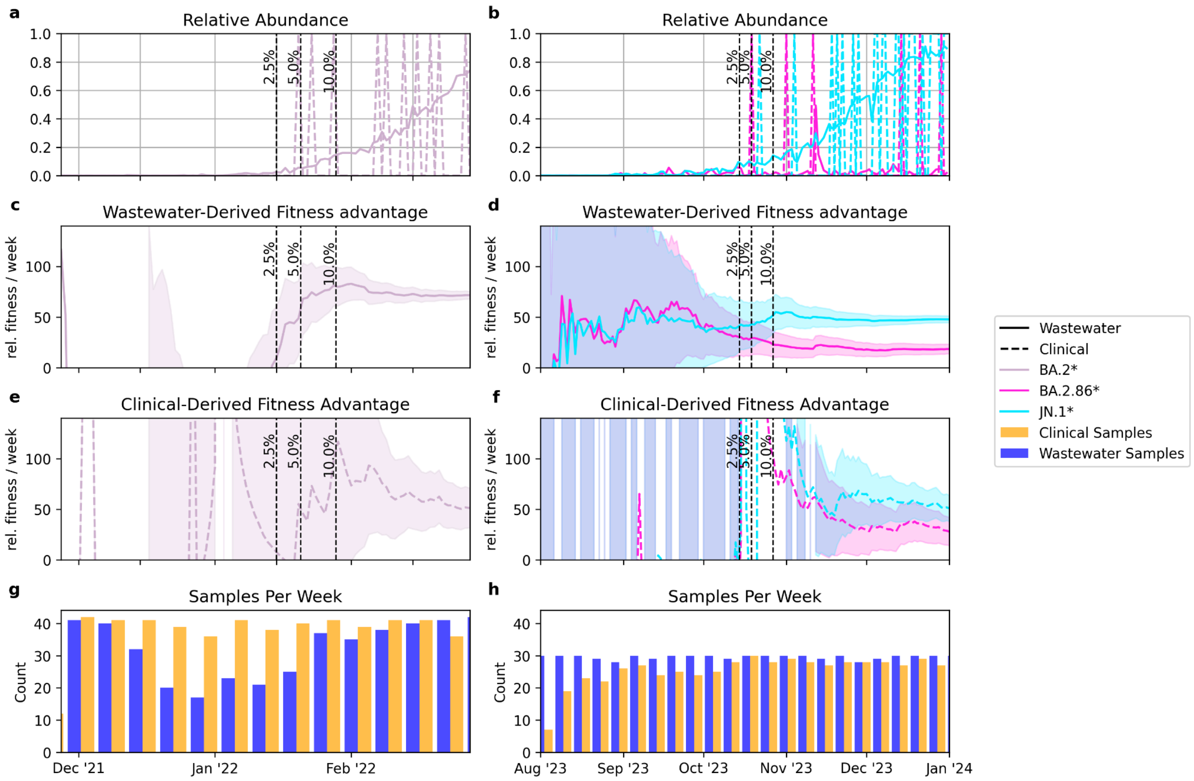


**Supplementary Fig. 1** Early estimation of the fitness advantage of SARS-CoV-2 variants using *Covvfit*, with subsampled clinical data. The clinical data here has been subsampled to a maximum of 1 sample per date, per location, to better match the sample size of the wastewater data. **a** Introduction and spread of the BA.2 variant, based on relative abundance estimates from wastewater (solid line) and clinical (dashed line) sequencing data. For illustration, the time series is shown for the WWTP of Zurich and its catchment population. The vertical lines represent times at which BA.2 is estimated to cross the 2.5%, 5% and 10% relative abundance thresholds in most of the surveyed WWTPs. **b** Similar time series illustrating the introduction of the BA.2.86 and JN.1 variants. **c,d** Estimates of fitness advantage (lines) and 95% confidence intervals (shaded bands), computed in a joint model of 6 WWTPs using data available up to sequential timepoints. The fitness advantage for BA.2 is relative (in percent) to BA.1, and the fitness advantage for BA.2.86 and JN.1 is relative to EG.5. **e,f** Similar sequential estimation of fitness advantage, based on the clinical data available in the regions in which the WWTPs are located, subsampled to a maximum of 1 sequence per day per location. **g,h** Number of samples per week used for the clinical data (blue) and for the wastewater data (orange).

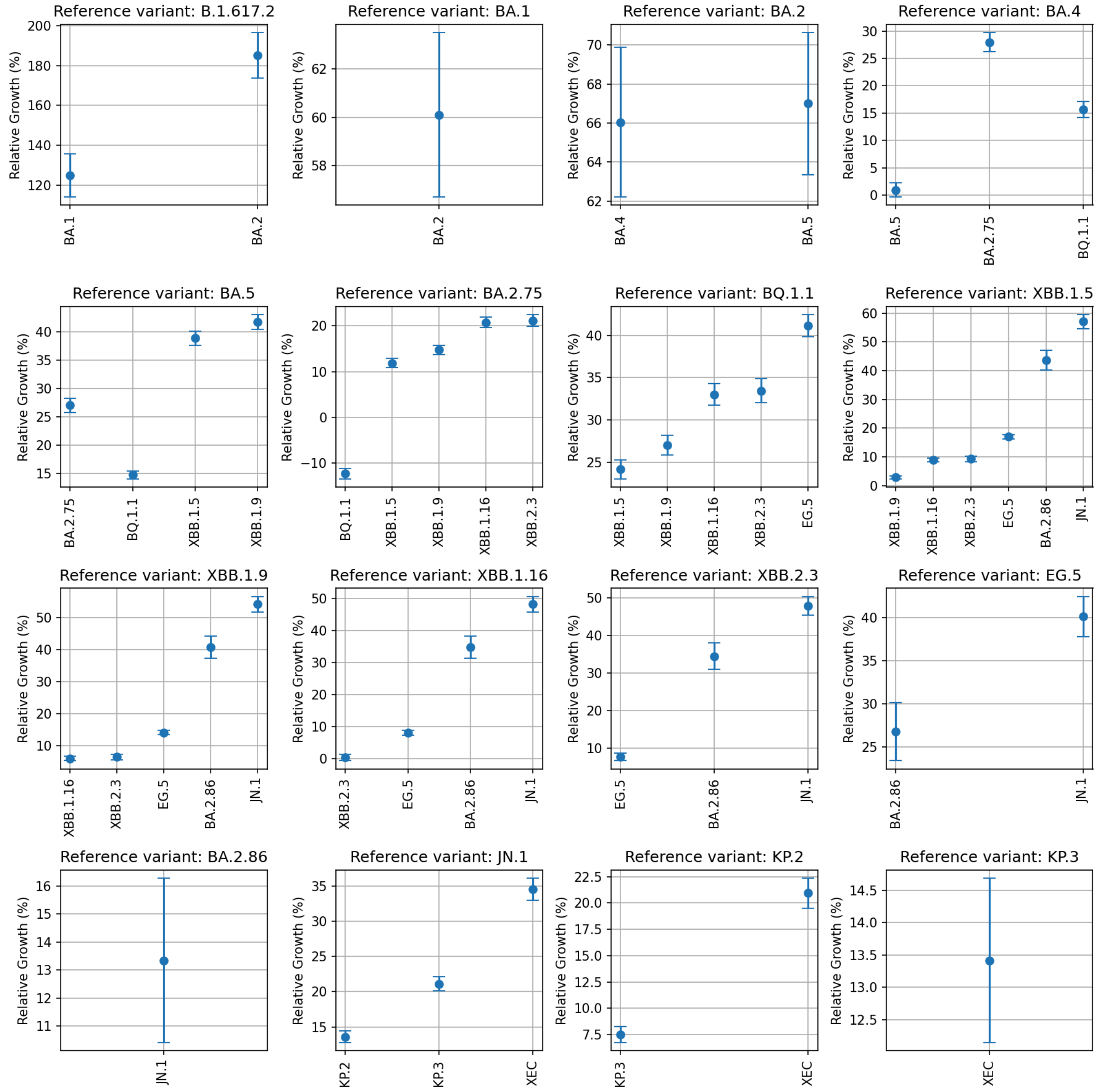


**Supplementary Fig. 2** Estimated continuous-time model fitness advantage of variants relative to reference variants they have been observed in competition with. The fitness advantage is calculated with a generation interval time of 7 days. Error bars represent 95% confidence intervals.

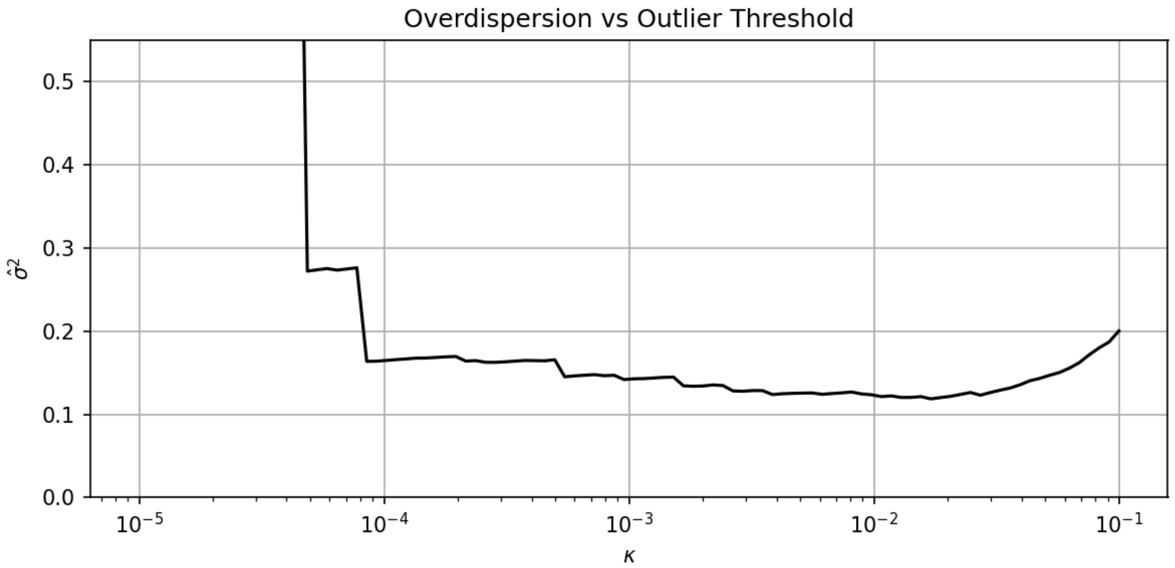


**Supplementary Fig. 3** Sensitivity analysis of the estimated overdispersion to the outlier threshold used in the calculation of the generalised Pearson residuals. The analysis shows that the overdispersion plug-in estimate $\hat{\sigma}^2$ is stable for thresholds $\kappa \in [10^{-4}, 10^{-2}]$.

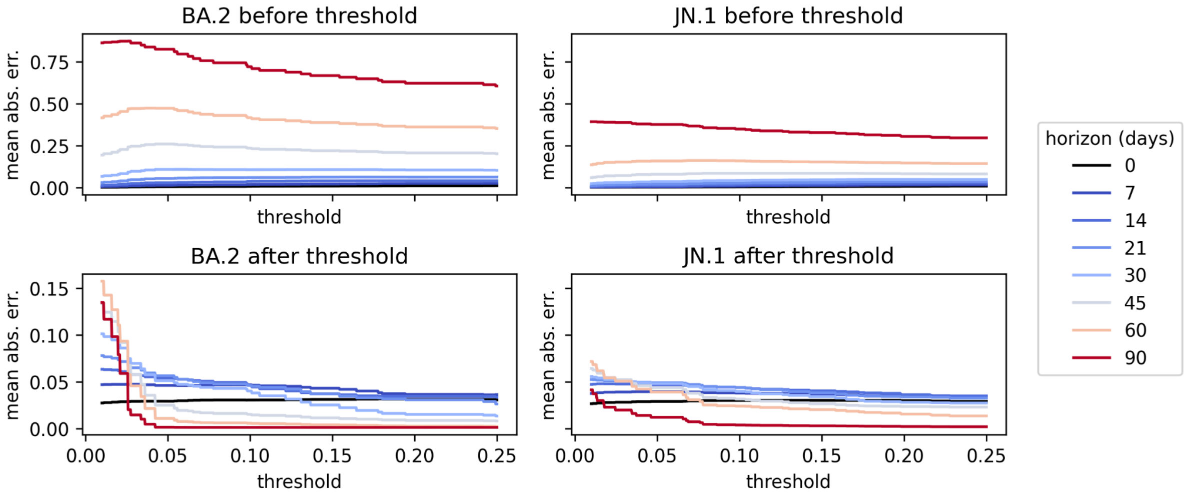


**Supplementary Fig. 4** Mean absolute prediction error for BA.2 and JN.1, averaged before or after the variant has reached a threshold estimated relative abundance in most surveyed cities, for varying threshold and forecast horizon.

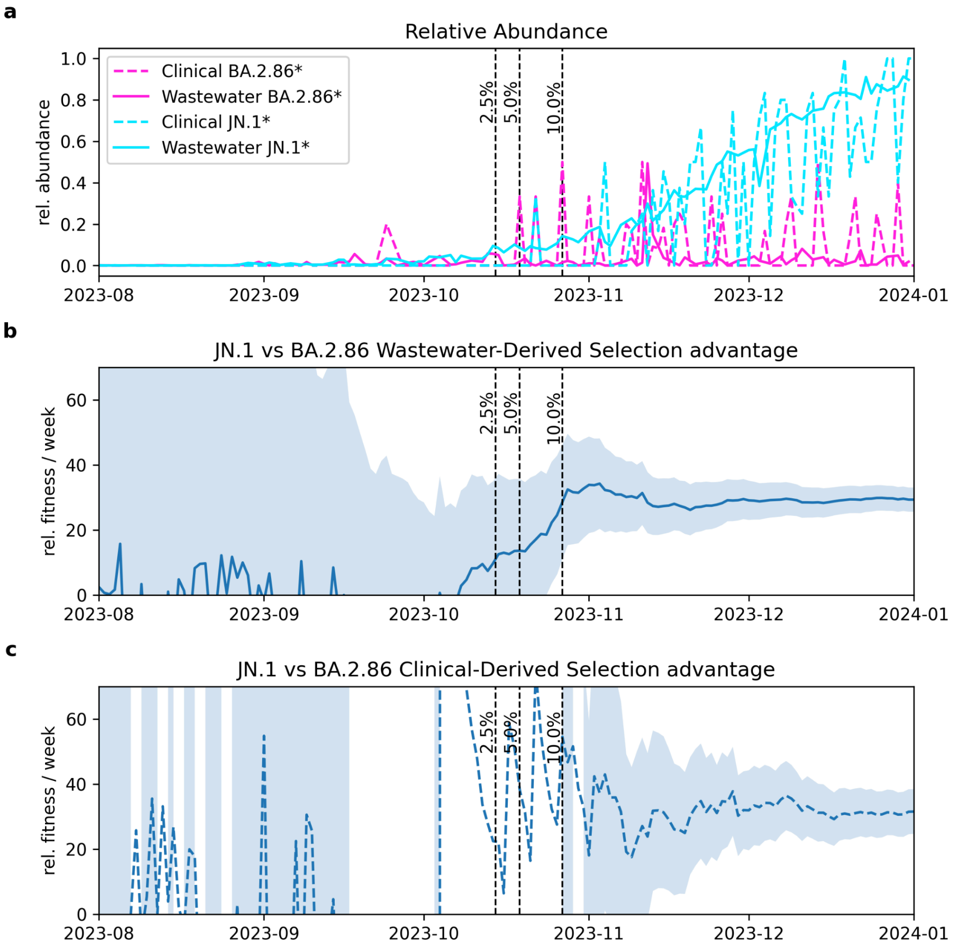


**Supplementary Fig. 5** Early estimation of the fitness advantage of JN.1 relative to BA.2.86. **a** Introduction of the BA.2.86 and JN.1 variants, illustrated by the wastewater data from Zurich. **b** Estimates of fitness advantage (lines) and 95% confidence intervals (shaded bands), computed in a joint model of 6 WWTPs using data available up to sequential timepoints. **c** Similar sequential estimates of fitness based on the clinical data.
